# Re-weighting *MC1R, ASIP* and *IRF4* risk variants optimises polygenic risk scores for keratinocyte cancer stratification in solid organ transplant recipients

**DOI:** 10.1101/2023.02.17.23286114

**Authors:** Mathias Seviiri, Matthew H. Law, Catherine M. Olsen, David C. Whiteman, Adele C. Green, Stuart MacGregor

**Author notes:** corresponding author- **Mathias Seviiri**; **Twitter: @MathiasSeviiri**, Statistical Genetics Lab, QIMR Berghofer Medical Research Institute, 300 Herston Road, Herston QLD 4006, Australia.

## Abstract

**Introduction:** Solid organ transplant recipients (SOTRs) are at much higher risk of developing squamous cell carcinoma (SCC) and basal cell carcinoma (BCC), compared to the general population. Previous studies have derived genetics-based predictors (polygenic risk scores, PRS) of SCC and BCC risk in SOTRs by assuming that genetic risk variants act in the same way in the general population as in SOTRs, but this assumption has not been fully tested.

**Objective:** To investigate whether known genetic risk variants for SCC and BCC have different effect sizes in SOTRs versus in non-transplantees, and if a re-weighted PRS would improve risk prediction.

**Methods:** We conducted genome-wide association studies for SCC and BCC separately in the non-transplant general population and in SOTRs, and compared the risks associated with selected common genetic variants for KC risk in SOTR vs non-transplant individuals from the UK Biobank. For regions with an increased log odds ratio in SOTRs, PRSs including these weights were validated in the QSkin study, and applied to the Australian STAR SOTR cohort.

**Results:** Effect sizes for functional variants in MC1R (rs1805007), *ASIP* (rs6059655), and *IRF4* (rs12203592) were much more strongly associated with the risk of KC in SOTRs than in non-transplantees. The proportional increase in the effect sizes ranged from 1.9-fold for rs6059655 and BCC risk (SOTRs log (OR)=0.49, 95%CI=0.00-0.98 vs log (OR)=0.26, 95%CI=0.24-0.30 in non-transplantees) to as high as 4.8-fold for rs1805007 and SCC risk (SOTR log (OR)=0.88, 95% CI=0.41-1.35 vs log (OR)=0.18, 95% CI=0.12-0.24 in non-transplantees). PRS with SOTR derived weights for these SNPs showed improved SCC/BCC risk stratification in the STAR Cohort, with the optimised PRS reclassifying 19% of SCC cases vs 8% using the standard PRS, and 18% of BCC cases vs 12% using the standard PRS.

**Conclusion:** Effect sizes for SCC and BCC risk for genetic variants in the *MC1R, ASIP and IRF4* genes are elevated in SOTRs, and correctly weighting these variants improves risk stratification based on polygenic risk.

## INTRODUCTION

Solid organ transplant recipients (SOTRs) have up to 100-fold and 20-fold increased risk for squamous cell carcinoma (SCC) and basal cell carcinoma (BCC), respectively compared to the general population ^1^. In addition, the majority (57%) of SOTRs develop multiple skin cancers ^2^. Studies have demonstrated that advanced age, male sex, fair skin, excessive ultraviolet radiation (UV) damage, red and blonde hair are significant predictors of SCC and BCC risk ^3^. This ultra-high risk in SOTRs has been mainly attributed to immunosuppression as a result of graft anti-rejection medication ^4–6^.

Genome-wide association studies (GWAS) have identified several germline genetic factors influencing BCC and SCC susceptibility in the general population ^7–10^. Genomic regions including *MC1R, IRF4, CDKN2A, HLA* genes, and *ASIP* have well known functions in influencing SCC and BCC susceptibility in the general population ^7–10^. Many variants, including those in *MC1R* and near *ASIP* likely modulate risk through their effects on pigmentation; others, like *IRF4* may act through multiple pathways including pigmentation, nevus count and immune-regulatory effects ^7,8^. To date no studies have assessed whether genetic risk variants have the same impact on risk in immunosuppressed individuals as they do in the general population.

SCC and BCC polygenic risk scores (PRSs) derived from GWASs in the general non-transplantee population have effectively stratified the SCC and BCC risk following immunosuppression in SOTRs ^11–14^. Ideally, a PRS for use in SOTRs would be derived from a SOTR training set but in practice the sample sizes of SOTRs will always be too small to allow derivation of accurate PRS weights, especially for single nucleotide polymorphism (SNPs) of moderate or small effect. However, if a subset of (large effect) variants have different effects in SOTRs relative to the general population, then PRS with weights derived from SOTR populations may better stratify risk of skin cancer in SOTRs. Therefore, this study sought to investigate whether existing well characterised genomic risk regions for SCC and BCC have different effect sizes in SOTR populations.

## METHODS

### Discovery cohort: UK Biobank

The UK Biobank (UKB) is a large (N∼ 500,000) population-based cohort of middle-aged participants (40-69 years) at the time of recruitment (2006-2010). Full details on participant recruitment, data collection and sample processing are published elsewhere ^15,16^. Phenotypic and genetic data were collected at baseline recruitment, and participants were followed up for disease outcomes including SCC and BCC. Both SCC and BCC were ascertained through participant data linkage with the National Cancer Registries in the UK. SCC and BCC statuses were based on the international classification of diseases (ICD) codes 10 (data field 40006), ICD 9 (data field 40013) and histological confirmation (data field 40011). Other skin cancer risk factor phenotypes used included; skin colour (data field 1747), tanning (data field 1727), childhood sunburns (data field 1737), and hair colour (data field 1747). Participants were genotyped using UKB Axiom Array and the UK BiLEVE Axiom Array, whilst imputation was done using the UK10K reference and Haplotype Reference Consortium panels ^16^. Full details on the quality control procedures for cleaning the genetic data have been published elsewhere ^16^. The National North West Multi-Centre Research Ethics Committee in the UK provided the ethical oversight over the UKB, and all participants provided written informed consent.

In this present study, we split the UKB dataset into two; the non-transplant population and the solid organ transplant recipient samples. First, the non-transplant dataset for SCC analysis included 7,402 SCC cases and 286,892 controls, while for BCC it comprised 20,791 cases and 286,893 controls. All participants were of European ancestry. Organ transplant recipients and those related to them (identity by descent pihat > 0.1) were excluded.

Second, for the organ transplant recipient dataset participants of European ancestry were selected based on transplantation and immunosuppressive medication use, as published before ^12^. This resulted in 112 SCC cases and 530 controls, and 100 BCC cases and 530 controls. In both the non-transplantee and transplantee populations, controls had no history of any cancer.

### Validation cohort: The QSkin Sun and Health Study (QSkin)

QSkin is a prospective cohort (N ∼ 43,000) of middle-aged participants at baseline recruited in Queensland. Details on the recruitment process, phenotypes collected etc are published elsewhere ^17^. About 18,000 participants also provided genetic data, which were genotyped using the Illumina Global Screening Array (Illumina, San Diego, USA). Participants with high genotype missingness (>3%), ancestry diversity from European population (6 SDs from the mean of PC1 and PC2 of 1000 Genomes European sample) were excluded, whilst SNPs with a low minor allele frequency (< 1%), a low GenTrain score (< 0.6) and Hardy-Weinberg equilibrium violation (P < 1 × 10^−6^) were removed. Genetic data were imputed using the Haplotype Reference Consortium panel ^18^.

Following recruitment, participants were followed up for SCC and BCC outcomes through both clinical confirmation and data linkage to the Australian pathology registers and the Medicare Australia database ^17^. Participants provided written informed consent, and QIMR Berghofer Medical Research Institute provided the ethical oversight to the study. This present study included 853 SCC cases and 5240 controls, and 2,064 BCC cases and 5240 controls. Controls had no history of any cancer or actinic keratoses.

### Testing Cohort: The Skin Tumours in Allograft Recipients (STAR) Cohort

The STAR cohort is a prospective study in Queensland, Australia comprising 600 SOTRs recruited between 2012 and 2014. Participants were recruited from two tertiary hospitals for transplant services; Prince Charles Hospital for lung transplant recipients and Princess Alexandra Hospital for kidney and liver transplant recipients. Details on recruitment, data collection, processing and participants follow-up have been published elsewhere and summarised here ^19–21^. At recruitment, information on socio-demographics, transplant (type of and age at transplantation), immunosuppression (duration, and type of immunosuppressive medication), skin colour, sun exposure, skin reaction to the sun, history of skin cancer and number of lifetime painful burns. In addition, biological samples were collected for genetic data.

375 participants including 30 liver, 93 lung and 252 kidney transplant recipients had their DNA extracted and genotyped using an Illumina Global Screening Array. Based on genetic data, individuals were excluded based on high genotype missingness (> 3%), relatedness (identity by descent pihat > 0.2), ancestry diversity from European samples in the HapMap phase 3 (> 6 standard deviations from the European mean of PC1 and PC2), and sex-mismatch ^13^. In addition, SNPs were removed based on a low call rate (< 95%), low MAF (< 1%), and Hardy-Weinberg equilibrium *P* < 1 × 10^−6 13^. Imputation to the Haplotype Reference Consortium panel ^18^ using the Michigan Imputation Server ^22^.

Participants were followed up until June 2016 to ascertain skin cancer outcomes (BCC and SCC). Incident skin cancer lesions were both clinically and histo-pathologically confirmed by dermatologist, physicians and through pathology registers. The human research ethics committees at the Metro South Hospital and Health Service, Brisbane, Australia and at QIMR Berghofer Medical Research Institute, Brisbane provided ethical oversight to the study. All participants provided written informed consent to participate in the study.

### Statistical analyses

#### Step 1: SCC and BCC GWAS in UKB (non-transplant and SOTRs)

We conducted case-control genome-wide association studies (GWAS) for SCC (7,402 cases and 286,892 controls), and BCC (20,791 cases and 286,893 controls) for the non-transplant participants of European ancestry in the UKB using SAIGE ^23^. We adjusted for age, sex, and population stratification using the first ten principal components (PCs). Analysis was restricted to SNPs with minor allele frequency of greater than 1%. In a similar way, using data for only SOTRs of European descent in the UKB we conducted another set of case-control GWAS for SCC (118 cases and 542 controls) and BCC (103 cases and 542 controls).

#### Step 2: Comparing effect estimates of selected common genetic variants in both transplant and non-transplant populations

The goal was to assess a small number of SNPs if the effect size in SOTRs was different to that in the general population; this would allow the computation of a more accurate PRS for use in predicting skin cancer in SOTRs. We selected top 10 genome-wide significant SNPs from the largest and latest meta-analyses for BCC and SCC ^10^. The SNPs selected have also been previously confirmed in other GWASs ^7,8,24^. We compared the risk of SCC and BCC associated with each SNP in SOTRs vs non-transplant individuals from the UK Biobank using the GWAS summary data above. We compared the effect sizes (log odds ratio) for risk variants rs1126809 (*TYR*), rs1805007 (*MC1R*), rs7528427 (*RCC2*), rs12070203 (*RHOU*), rs214803 (*TGM3*), rs62209647 (*RALY*), rs6059655 (*ASIP*), rs35407 (*SLC45A2*), rs12203592 (*IRF4*), and rs10810657 (*BNC2*) (**Table 1**), in SOTRs versus in non-transplant population for both SCC and BCC. rs6059655 and rs62209647 are in high LD (r^2^=0.93) in Europeans so we selected only one of them (rs6059655). Although rs35407 showed increased effect for SCC, it had a big standard error and wide confidence interval, and it showed no increase in another independent sample of SOTRs (**Table 1**). Thus, it was not selected in the final model. In addition to its effect being increased in the UKB SOTRs, rs6059655 was significantly increased in another independent cohort of SOTRs (STAR; increase: 3.24-fold, P= 6.4 × 10^−3^) so it was selected for BCC.

**Table 1.**
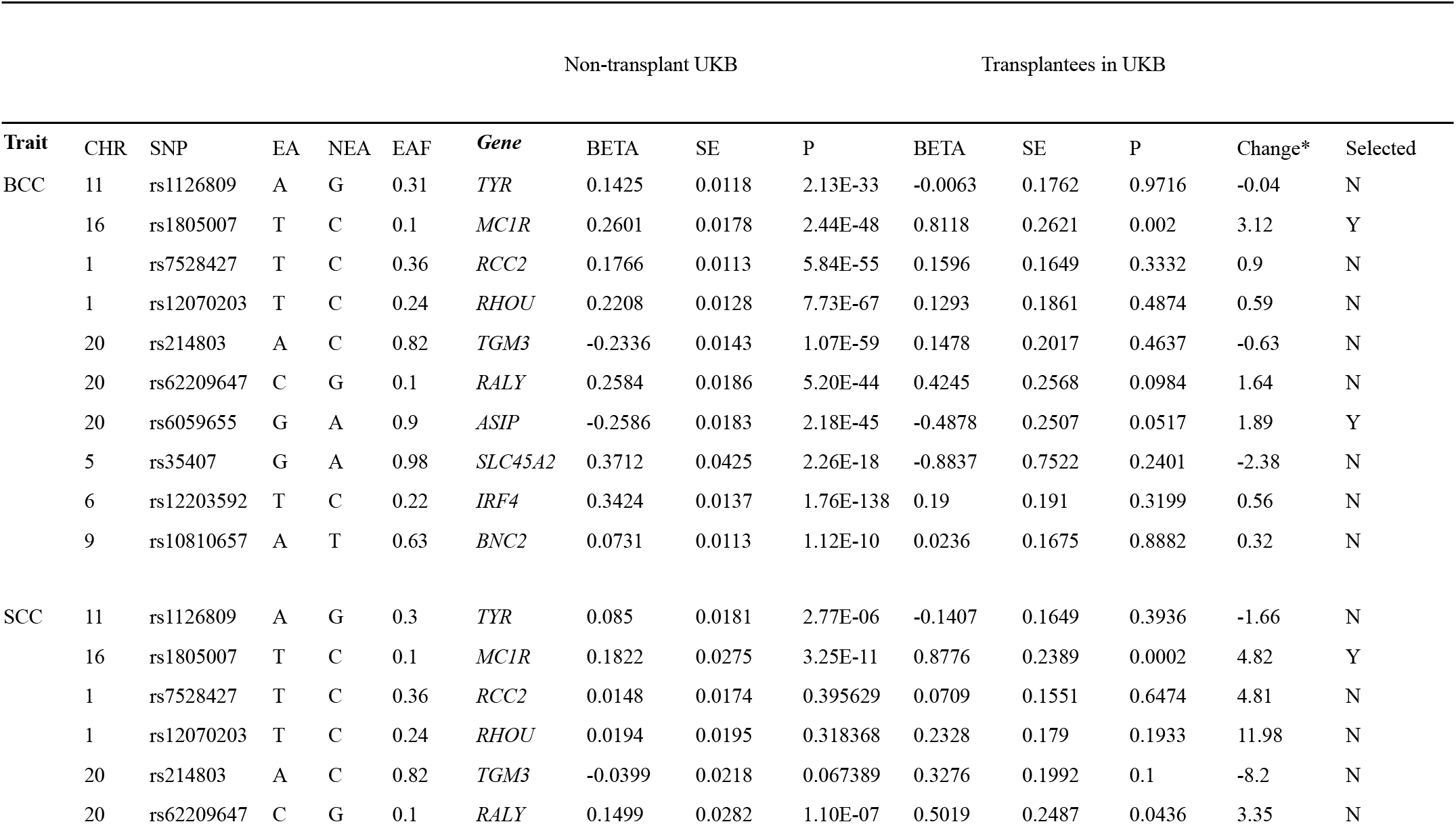
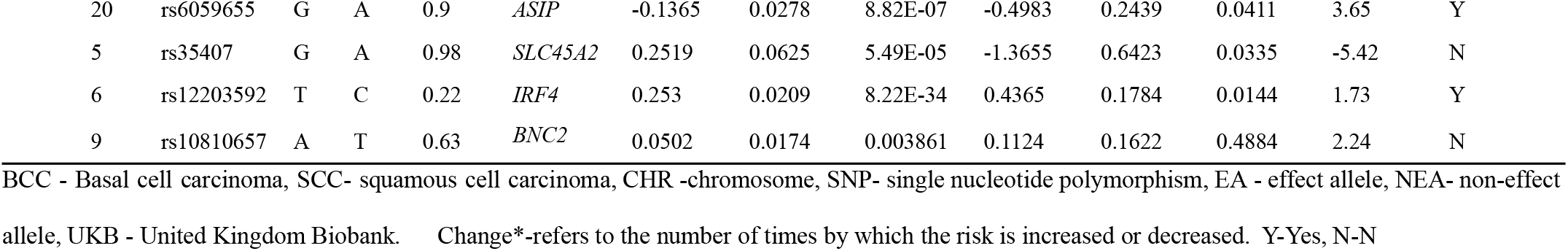
Common risk genetic variants for KC and their associated risk in the non-transplant and transplant population.

#### Step 3: Construction of a standard PRS, PRS validation in the QSkin cohort and model selection

Using the non-transplant GWAS data for BCC and SCC, we constructed several PRS models with independent SNPs using a varying P Value threshold (S1: P < 5 × 10^−8^; S2: P < 10^−7^; S3: P < 10^−6^; S4: P < 10^−5^; S5: P < 10^−4^; S6: P < 10^−3^; S7: P < 10^−2^; S8: P < 10^−1^). Next we validated the models in an independent external cohort; QSkin to select the best model based on a model-fit parameter, Nagelkerke’s R^2 25^ calculated using the predictABEL R package ^26^. Analysis was done separately for SCC and BCC.

#### Step 4: Generation of optimised BCC and SCC PRS using OTR weights for genomic regions with increased risk

Using the validated PRS models generated for the standard PRSs above, we generated a new optimised PRS for BCC and (separately) for SCC, accounting for the genetic risk differences for OTRs vs general population. For each of the 10 SNPs noted in step 2, we selected SNPs for re-weighting by examining if the log OR was significantly different (P < 0.05) between the OTR and general population groups. For regions with an increased log odds ratio for KC, PRSs were derived using weights from UK Biobank OTRs instead of using the UKB non-transplant population weights. That is, for SNP rs1805007 for *MC1R* (BCC and SCC), rs6059655 for (*ASIP*) (BCC and SCC), and rs12203592 for *IRF4* (only SCC), we replaced the log odds ratio estimate used as a weight in the PRS with the estimate from the OTR set. For the rest of the SNPs, there was no strong evidence for a different effect in OTRs vs general population and weights from the UKB non-transplantees were used.

#### Step 5: Comparing model performances in STAR (Optimised vs Standard PRS)

We applied the standard PRSs (from Step 3) and the optimised PRSs (from Step 4) to an independent set of OTRs from the STAR Cohort. We evaluated and compared model performance for both versions (Optimised vs Standard PRS). First, we compared the magnitude of their association with BCC or SCC risk. E.g. the OR per SD increase in the PRS for BCC (BCC Optimised PRS OR vs BCC Standard PRS OR).

Second, we used the net reclassification improvement index based on the predictABEL R package ^26^, to assess the number of people reclassified to an appropriate risk group (tertile) after adding the PRS to the traditional risk factor models. This was done for both the standard and optimised PRSs. Next, we compared how well the optimised PRS reclassified OTRs compared to standard PRS for both BCC and SCC separately.

## 1.1 RESULTS

### Summary statistic for the STAR participants

Descriptive information for the STAR participants is presented in **Supplementary 1**. We restricted this analysis to 337 participants for the SCC analysis and 331 for BCC. Participants were generally middle aged with a mean (SD) age of 44.4 (14.2) years at the first transplantation, whilst the majority (65.6%) were male. After three years of follow-up, 36.8% and 35.6% developed SCC and BCC respectively.

### Genomic regions with increased risk of SCC and BCC in organ transplant recipients

The magnitude of the effect of the lead SNPs in the *MC1R, ASIP* and *IRF4* were increased by 4.82-fold, 3.65-fold and 1.73-fold respectively, for SCC risk in transplantees compared to non-transplantees (**Figure 1**). Similarly, for BCC risk the magnitude of the effect of the lead SNPs in *MC1R* and *ASIP* were increased by 3.12-fold and 1.89-fold respectively (**Figure 2**). Effect sizes for variants in *TYR, RCC2, RHOU, TGM3* and *BNC2 regions* were not significantly elevated or reduced in SOTRs. Therefore, we used the larger effect sizes for *MC1R* and *ASIP* to derive optimised PRSs for BCC, and *MC1R, ASIP* and *IRF4* for SCC.

**Figure 1.**
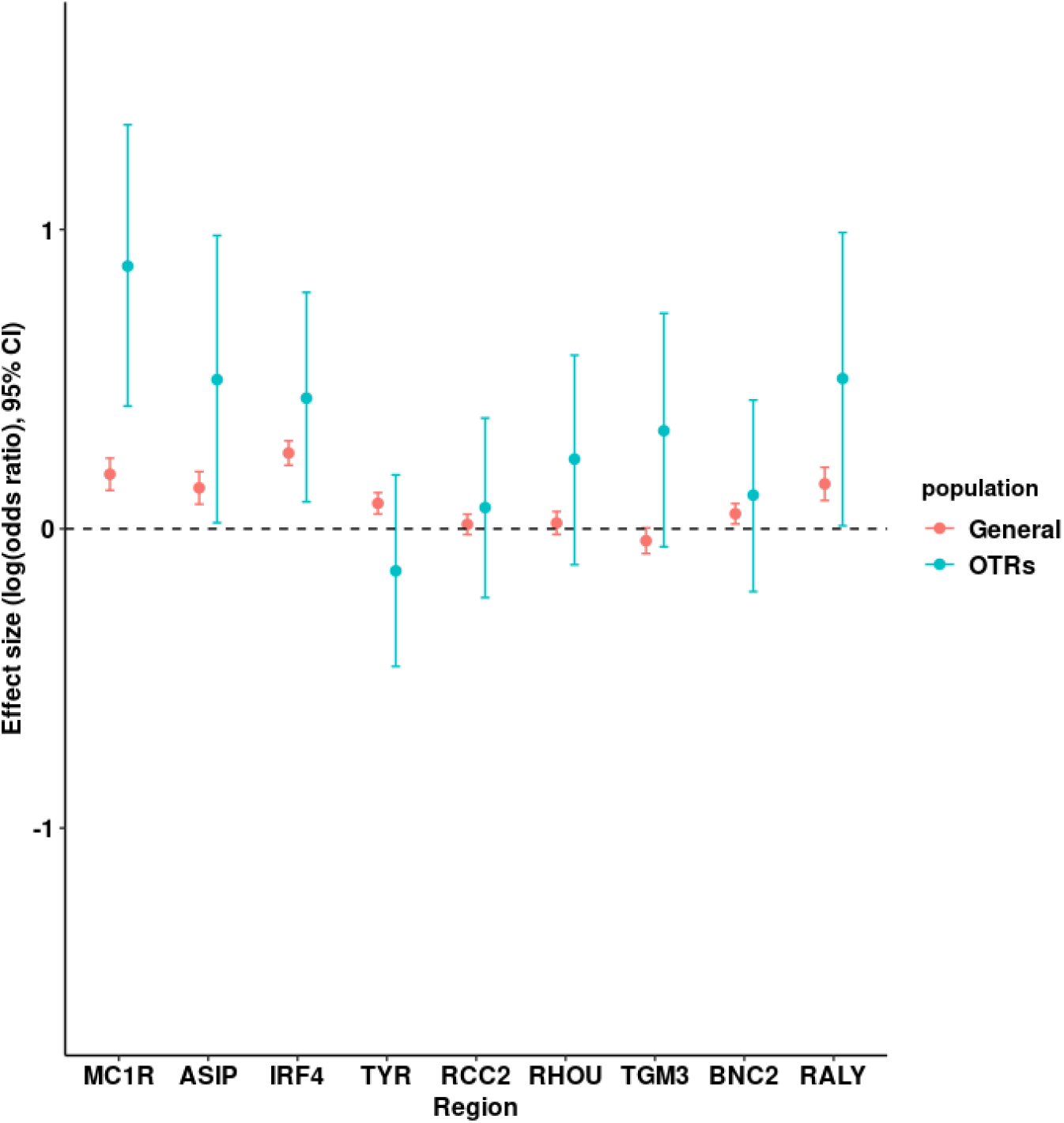
Comparison of the weights (log (odds ratios) for the common genetic risk variants for KC in the transplantees versus non-transplantees for SCC risk. The x-axis represents the genomic regions, whilst the y-axis represents effect in log odds ratio for the selected genetic variants in each of the genomic regions, for both organ transplant recipients (cyan) and non-transplantee (red) in the UK Biobank. OTRs - organ transplant recipients.

**Figure 2.**
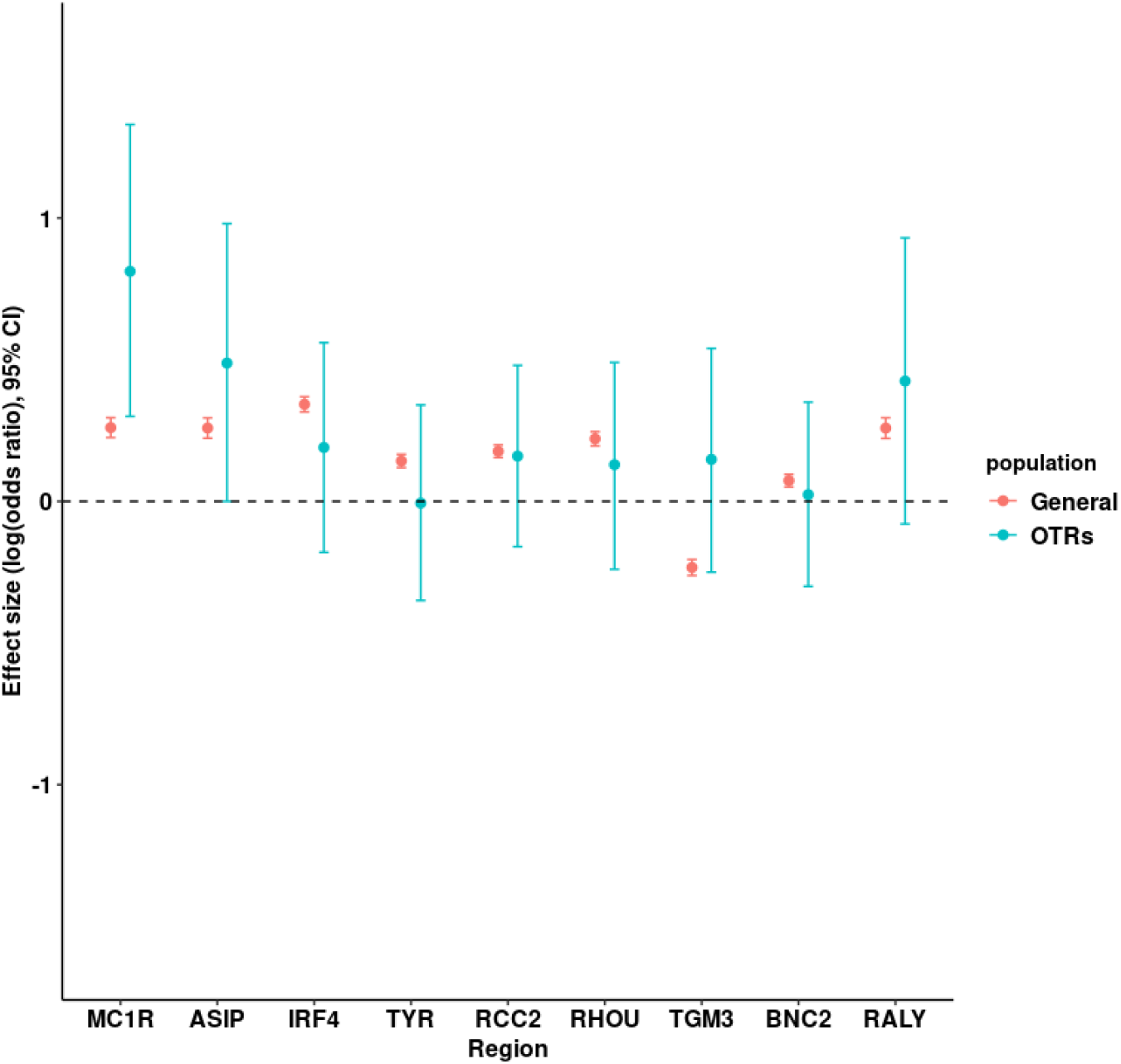
Comparison of the weights (log (odds ratios) for the common genetic risk variants for KC in the transplantees versus non-transplantees for BCC risk. The x-axis represents the genomic regions, whilst the y-axis represents effect in log odds ratio for the selected genetic variants in each of the genomic regions, for both organ transplant recipients (cyan) and non-transplantee (red) in the UK Biobank. The error bars represent the 95% confidence intervals for log odds ratios. OTRs - organ transplant recipients

### The best PRS performing models in the QSkin validation cohort

The PRS model S3 (P < 10^−6^) with a Nagelkerke’s R^2^ of 25.7 % was the best performing model for SCC, and S4 (P < 10^−5^; Nagelkerke’s R^2^ =24.0%) for BCC (**Figure 3**). In the subsequent analyses in the STAR cohort only the S3 (SCC) and S4 (BCC) PRS models were used.

**Figure 3.**
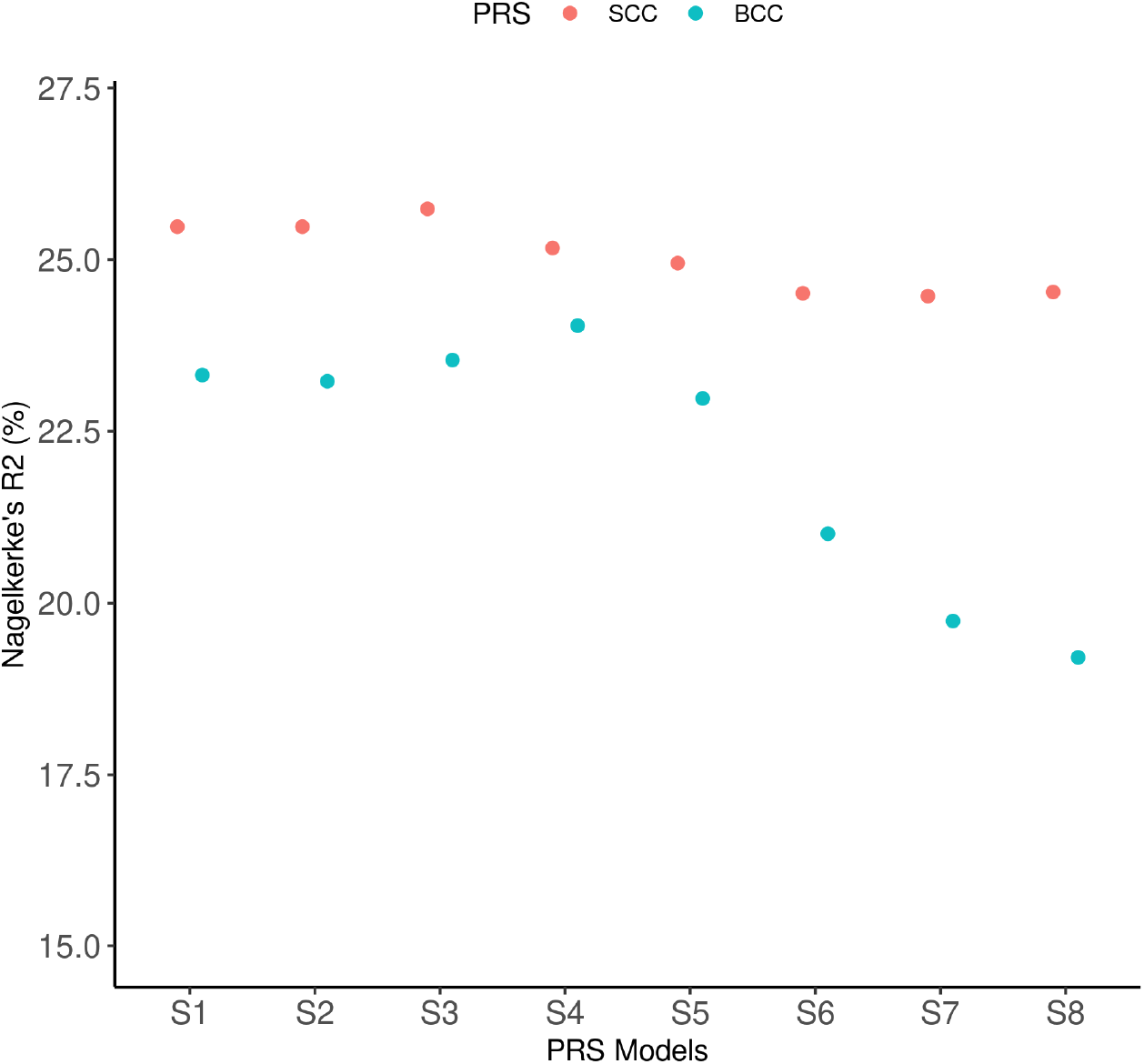
The performance of SCC and BCC PRS prediction models in the validation cohort. The x-axis represents the prediction models at varying P-value thresholds; S1: P < 5 × 10^−8^, S2: P < 10^−7^, S3: P < 10^−6^, S4: P < 10^−5^, S5: P < 10^−4^, S6: P < 10^−3^, S7: P < 10^−2^, and S8: P < 10^−1^. The y-axis represents Nagelkerke’s R^2^ (%) for each of the prediction models. The red and cyan colour represent the SCC and BCC PRSs respectively. The models with the highest Nagelkerke’s R^2^ (S3 for SCC and S4 for BCC) represent the best predictive models.

### Comparison of the association with KC risk for optimised versus standard PRS in the STAR Cohort

After adjusting for the established risk factors and the first 10 PCs, the risk of SCC per unit increase in PRS increased by 55% for the optimised SCC PRS (OR per SD = 1.55, 95% CI = 1.17 - 2.06, P= 2.4 × 10^−3^), compared to the 27% for the standard SCC PRS (**Figure 4**). In a similar way, the risk of BCC was increased by 50% for the optimised BCC PRS (OR per SD = 1.50, 95% CI = 1.13 - 1.98, P= 4.6 × 10^−3^), compared to the 36% for the standard BCC PRS (OR per SD = 1.36, 95% CI = 1.04 - 1.79, P= 0.02396) (**Figure 4**).

**Figure 4.**
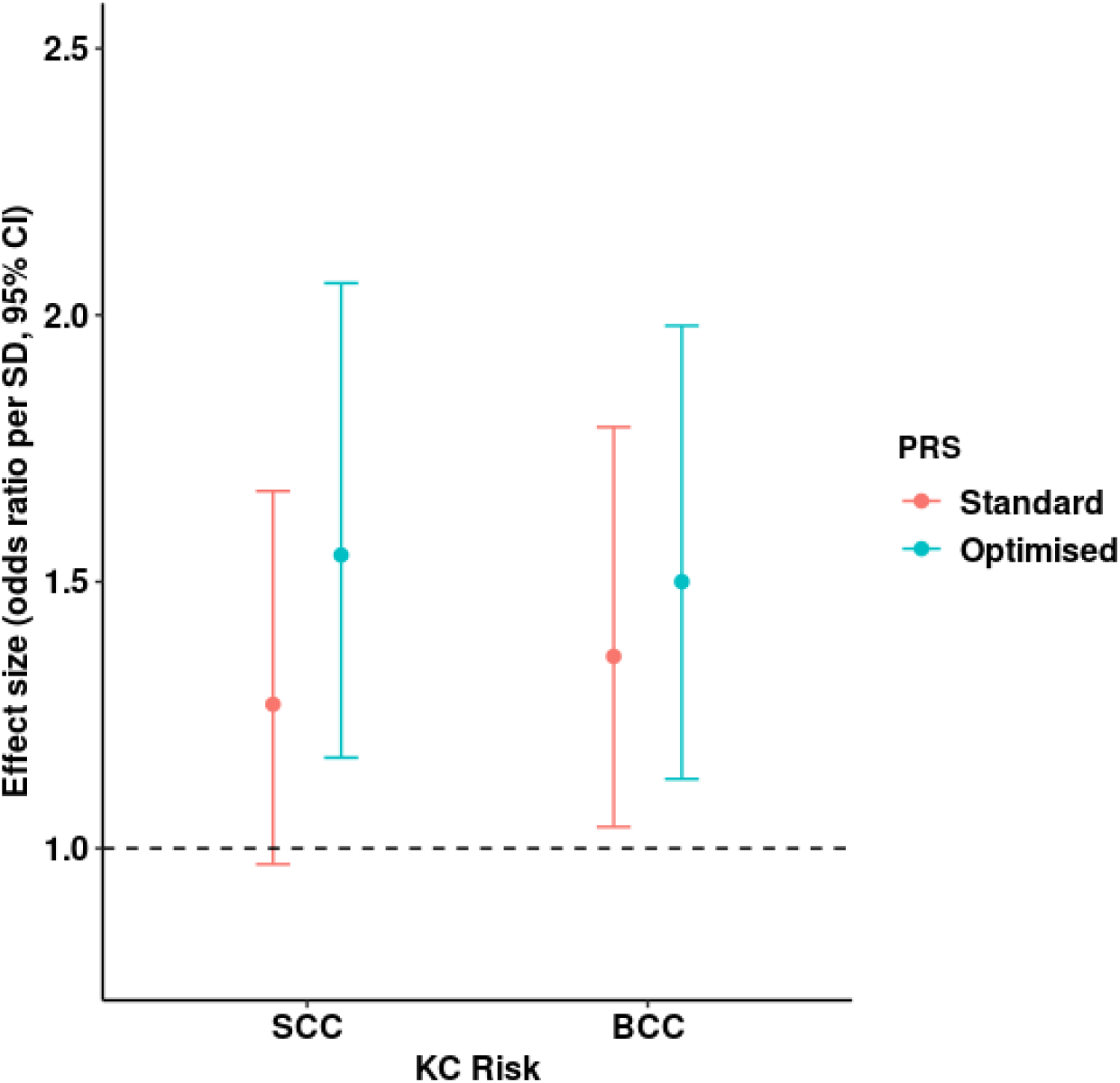
Comparison for the association of the optimised vs standard PRSs and the risks of BCC and SCC among SOTRs in the STAR cohort. The red and cyan colours represent the standard and optimised PRS respectively. The x-axis represents the two keratinocyte cancers; squamous cell carcinoma (SCC) and basal cell carcinoma (BCC). The y-axis shows the effect of the association in odds ratio per standard deviation increase in the PRS and the 95 % confidence intervals. The black dashed line shows a null effect. Logistic regression was used for analysis adjusting for established skin cancer risk factors (age, sex, skin colour and sun exposure and history of keratinocyte cancer) + 10 PCs.

### Comparison of KC risk reclassification improvement for optimised PRS versus standard PRS

Leveraging genetic variants in the *MC1R, ASIP* and *IRF4* genes, the SCC optimised PRS accurately re-stratified (after a clinical risk factor model) 29.20% and 12.61% OTRs with moderate and high genetic risk for SCC respectively (**Figure 5**). The standard SCC PRS was able to reclassify only about 12.39% and 6.31% for SCC risk in the moderate and high genetic risk strata respectively. Overall, the optimised SCC PRS outperformed the standard PRS in terms of accurately reclassifying individuals in their respective risk groups (Optimised PRS = 17.21% reclassified, versus standard PRS = 8.90% reclassified) (**Figure 5**). In a similar way, the optimised BCC PRS outperformed the standard PRS by reclassifying 32.82%, 10.91% and 16.62% participants in the moderate, high and overall categories for BCC risk respectively; compared to 25.45%, 8.18% and 14.20% in the respective categories by the standard PRS (**Figure 6**).

**Figure 5.**
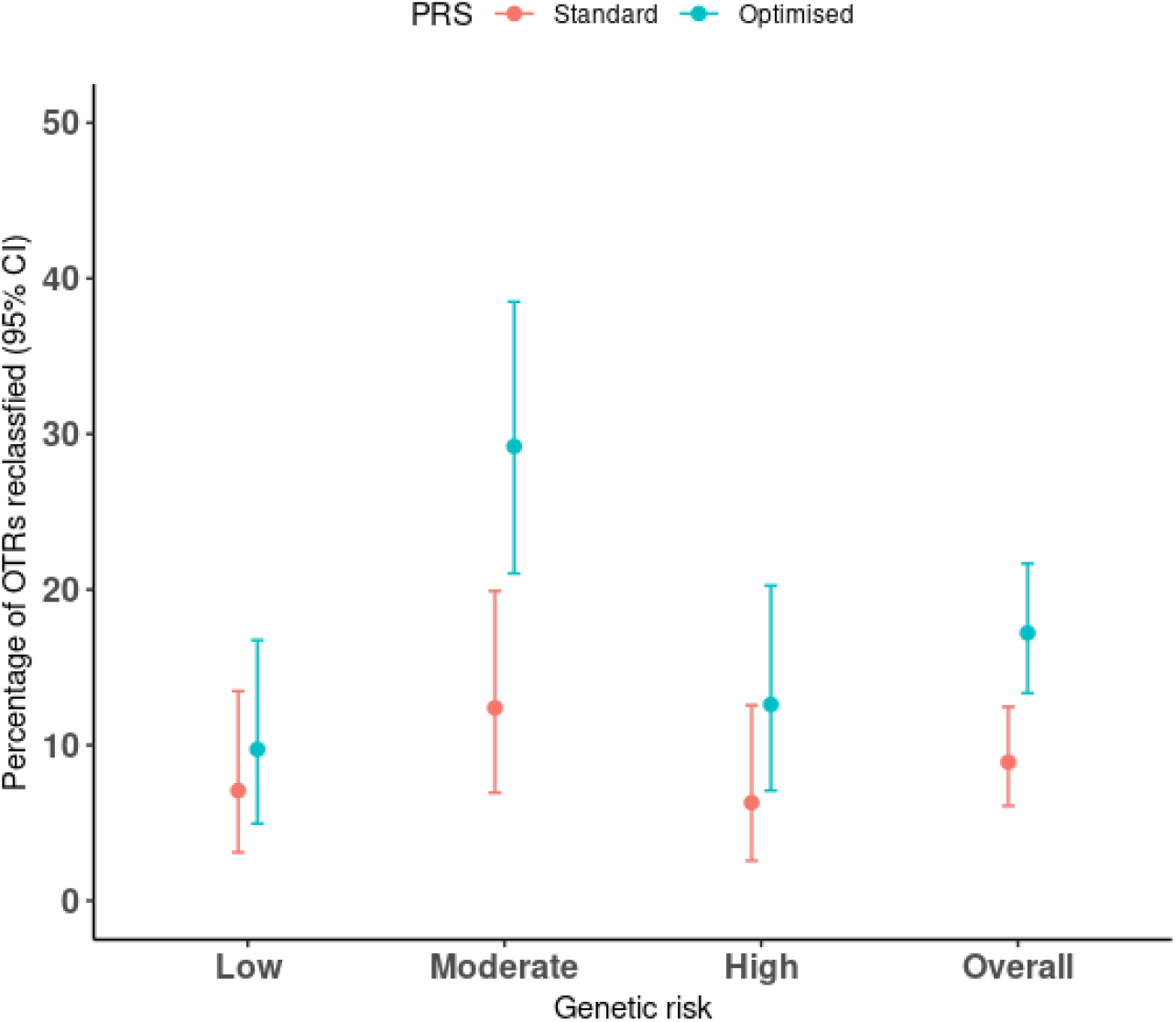
Comparison of the percentage of SOTRs reclassified after adding the optimised versus standard SCC PRS to a prediction model with only traditional skin cancer risk factors in the STAR cohort. The x-axis represents the genetic risk strata; low (bottom 20% PRS), moderate (middle 60%) and high (top 20%). The y-axis represents the percentage of the transplantees that got stratified within each risk stratum, and also overall in the cohort. The error bars show the 95% confidence intervals for the percentage. The red and cyan colours represent the standard and optimised PRS respectively.

**Figure 6.**
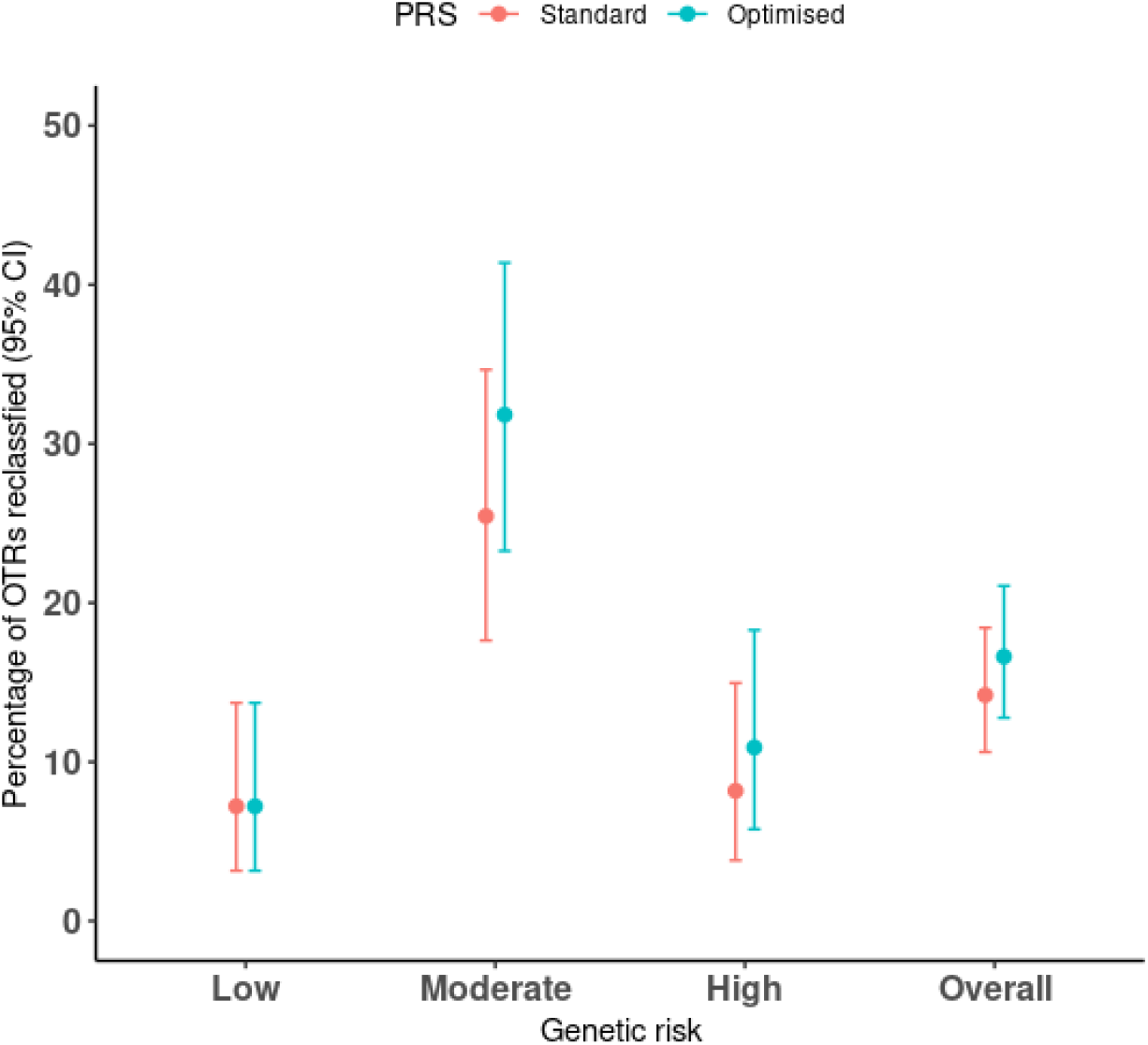
Comparison of the percentage of SOTRs reclassified after adding the optimised versus standard SCC PRS to a prediction model with only traditional skin cancer risk factors in the STAR cohort. The x-axis represents the genetic risk strata; low (bottom 20% PRS), moderate (middle 60%) and high (top 20%). The y-axis represents the percentage of the transplantees that got stratified within each risk stratum, and also overall in the cohort. The error bars show the 95% confidence intervals for the percentage. The red and cyan colours represent the standard and optimised PRS respectively.

## 1.2 DISCUSSION

We and others have shown that a KC PRS derived from the general population is able to predict risk in both high UV and low UV settings ^11–13^. In this paper we explored if there are differences in the magnitude of risk conferred by known germline genetic risk factors for OTRs vs general population in the UKB. We found large differences in the risk conferred by functional *MC1R, ASIP and IRF4* variants (5-fold, 4-fold, and 2-fold for SCC respectively), but this was in general not true for other KC risk variants e.g. in the *TYR*, and *RCC2*.

*MC1R, ASIP* and *IRF4 influence skin cancer risk through the pigmentation pathway*. For example, *MC1R* and *ASIP* play an important role in determining pigment variation by controlling the ratio of eumelanin (dark brown to black) and pheomelanin (light colour or fair skin, and red hair) ^27,28^. Pheomelanin is phototoxic, generating free radicals following UV radiation and leading to UV-induced skin damage whilst eumelanin is photoprotective ^29^. Individuals that only produced pheomelanin (fair skinned and or red hair individuals) are at a higher risk of developing skin cancer due to having no/less eumelanin, and some risk from the (photo)toxic role for pheomelanin. Our results imply that eumelanin and pheomelanin might alter the KC risk in SOTRs in a different way to in the general population. Therefore, this suggests that immunosuppression (SOTR status) negatively interacts with skin colour and UV radiation, increasing the KC risk more in people with less eumelanin. This is in keeping with previous observational studies which identified skin colour (fair-skinned vs non-fair skin OR: 8.18, 95%CI = 5.49-12.18), as a leading determinant of SCC in SOTRs compared to other risk factors like age (50+ vs < 50 years OR : 2.73, 95% CI = 2.24-3.34), sex (male vs female OR : 1.67, 95% CI = 1.4-2.01), and type of transplant (thoracic vs abdominal OR: 1.53, 95%CI = 1.27 -1.85) ^30^.

Compared to the standard PRS, the optimised PRS improved the risk stratification of KC in SOTRs. This was achieved by utilising both SNP weights (log odds ratio) from the non-transplant and SOTRs discovery GWAS, and upweighting the independent SNPs in the *MC1R* and *ASIP* genes selected for the PRS. E.g. in a well powered discovery SCC GWAS, the *MC1R, ASIP* and *IRF4* weights could be multiplied by a factor **F** which is equivalent to 4.82, 3.65 and 1.78 respectively before calculating the individual scores. In the case of BCC, the *MC1R* and *ASIP* SNP effect sizes could be multiplied by 3.12 and 1.89 respectively. Therefore, given that currently there are no large discovery KC GWAS in SOTRs to develop an SOTR specific PRS, PRS generated from the general non-transplant population should consider upweighting the *MC1R* and *ASIP* SNPs before applying them to SOTRs.

This study has a number of strengths. The BCC and SCC risk comparisons between the OTRs and non-transplantees were made in the same population source and UV environment (UKB). This minimised biases from environment and population stratification that would have occurred e.g. from one GWAS was conducted in the UK population and the other in Australia. In addition, the non-transplant GWASs were well-powered (SCC: 7,402 cases and 286,892 controls, and BCC: 20,791 cases and 286,893 controls) to enable us to make precise estimates for the genetic variants.

One caveat is the small sample size of SOTRs in the UKB to reliably detect other SNPs with higher effect sizes. This could imply that with the current modest sample size, only SNPs with a large effect size (e.g. in *MC1R, ASIP and IRF4*) can be reliably detected. It may be that in a suppressed immune system all variants have different effect sizes relative to the general population, but we did not have power for those with more modest effect sizes. Future studies that include larger sample sizes for SOTRs would be helpful. However, using data from an independent cohort of SOTRs in Australia we confirmed these elevated results in the *MC1R* and *ASIP*.

In conclusion, genetic variants in the *MC1R, ASIP* and *IRF4* genes make unusually large contributions to the elevated risk of SCC and BCC in SOTRs and offer improved KC risk stratification based on polygenic risk modelling. Genetic variants in pathways for very fair skin puts SOTRs at a much higher risk of SCC/BCC compared to people with non-fair skin complexion.

## Data Availability

The polygenic scores generated in this study will be accessed through the Polygenic Score Catalog (https://www.pgscatalog.org/). The underlying data for each cohort can be accessed through application to the UK Biobank (http://www.ukbiobank.ac.uk/wp-content/uploads/2012/09/Access-Procedures-2011-1.pdf), QSkin (Principal Investigator David Whiteman at), and STAR (Principal Investigator Adele Green at).

## Author Contributions

Conceptualization: MS, MHL, SM; Data curation: MS, MHL, SM; Formal analysis: MS; Funding acquisition: SM, MHL, DCW, ACG; Investigation: MS, MHL, DCW, CMO, ACG, SM; Methodology: MS, MHL, SM; Project administration: MS, MHL, SM; Resources: MHL, DCW, CMO, SM; Software: MS; Supervision: SM, MHL; Visualisation: MS; Writing - original draft preparation: MS. All authors contributed to and approved the final version of the manuscript.

## Acknowledgments

The study was supported by a program grant (APP1073898) and a project grant (APP1063061) from the Australian National Health and Medical Research Council (NHMRC). SM and DCW are supported by Research Fellowships from the NHMRC. MS was supported by the Australian Government Research Training Program (RTP) and the Faculty of Health Scholarship at Queensland University of Technology, Australia. This study was conducted using data from the UK Biobank (application number 25331), and the QSkin Sun and Health Study (Australia), and the STAR Cohort (Australia).

## REFERENCES

1. Lindelöf, B., Sigurgeirsson, B., Gäbel, H. & Stern, R. S. Incidence of skin cancer in 5356 patients following organ transplantation. Br. J. Dermatol. 143, 513–519 (2000).

2. Wehner, M. R. et al. Risks of Multiple Skin Cancers in Organ Transplant Recipients: A Cohort Study in 2 Administrative Data Sets. JAMA Dermatol. 157, 1447–1455 (2021).

3. Gordon, R. Skin cancer: an overview of epidemiology and risk factors. Semin. Oncol. Nurs. 29, 160–169 (2013).

4. Gerlini, G., Romagnoli, P. & Pimpinelli, N. Skin cancer and immunosuppression. Crit. Rev. Oncol. Hematol. 56, 127–136 (2005).

5. Thoms, K.-M. et al. Cyclosporin A, but not everolimus, inhibits DNA repair mediated by calcineurin: implications for tumorigenesis under immunosuppression. Experimental Dermatology vol. 20 232–236 Preprint at https://doi.org/10.1111/j.1600-0625.2010.01213.x (2011).

6. Guba, M., Graeb, C., Jauch, K.-W. & Geissler, E. K. Pro- and anti-cancer effects of immunosuppressive agents used in organ transplantation. Transplantation 77, 1777–1782 (2004).

7. Chahal, H. S. et al. Genome-wide association study identifies 14 novel risk alleles associated with basal cell carcinoma. Nat. Commun. 7, 12510 (2016).

8. Chahal, H. S. et al. Genome-wide association study identifies novel susceptibility loci for cutaneous squamous cell carcinoma. Nat. Commun. 7, 12048 (2016).

9. Seviiri, M. et al. A multi-phenotype analysis reveals 19 novel susceptibility loci for basal cell carcinoma and 15 for squamous cell carcinoma. Preprint at https://doi.org/10.1101/2022.03.06.22271725 (2022).

10. Liyanage, U. E. et al. Combined analysis of keratinocyte cancers identifies novel genome-wide loci. Hum. Mol. Genet. 28, 3148–3160 (2019).

11. Stapleton, C. P. et al. Polygenic risk score as a determinant of risk of non-melanoma skin cancer in a European-descent renal transplant cohort. Am. J. Transplant 19, 801–810 (2019).

12. Seviiri, M. et al. Polygenic risk scores allow risk stratification for keratinocyte cancer in organ transplant recipients. J. Invest. Dermatol. 141, 325–333.e6 (2021).

13. Seviiri, M. et al. Polygenic Risk Scores Stratify Keratinocyte Cancer Risk among Solid Organ Transplant Recipients with Chronic Immunosuppression in a High Ultraviolet Radiation Environment. J. Invest. Dermatol. 141, 2866–2875.e2 (2021).

14. Stapleton, C. P., Chang, B.-L., Keating, B. J., Conlon, P. J. & Cavalleri, G. L. Polygenic risk score of non-melanoma skin cancer predicts post-transplant skin cancer across multiple organ types. Clin. Transplant. 34, e13904 (2020).

15. Sudlow, C. et al. UK biobank: an open access resource for identifying the causes of a wide range of complex diseases of middle and old age. PLoS Med. 12, e1001779 (2015).

16. Bycroft, C. et al. The UK Biobank resource with deep phenotyping and genomic data. Nature 562, 203–209 (2018).

17. Olsen, C. M. et al. Cohort profile: the QSkin Sun and Health Study. Int. J. Epidemiol. 41, 929–929i (2012).

18. McCarthy, S. et al. A reference panel of 64,976 haplotypes for genotype imputation. Nat. Genet. 48, 1279–1283 (2016).

19. Iannacone, M. R. et al. Sun Protection Behavior in Organ Transplant Recipients in Queensland, Australia. Dermatology 231, 360–366 (2015).

20. Hartman, R. I., Green, A. C., Gordon, L. G. & Skin Tumours and Allograft Recipients (STAR) Study. Sun Protection Among Organ Transplant Recipients After Participation in a Skin Cancer Research Study. JAMA Dermatol. 154, 842–844 (2018).

21. Plasmeijer, E. I. et al. Extreme Incidence of Skin Cancer in Kidney and Liver Transplant Recipients Living with High Sun Exposure. Acta Derm. Venereol. 99, 929–930 (2019).

22. Das, S. et al. Next-generation genotype imputation service and methods. Nat. Genet. 48, 1284–1287 (2016).

23. Zhou, W. et al. Efficiently controlling for case-control imbalance and sample relatedness in large-scale genetic association studies. Nat. Genet. 50, 1335–1341 (2018).

24. Asgari, M. M. et al. Identification of Susceptibility Loci for Cutaneous Squamous Cell Carcinoma. J. Invest. Dermatol. 136, 930–937 (2016).

25. Nagelkerke, N. J. D. A note on a general definition of the coefficient of determination. Biometrika 78, 691–692 (1991).

26. Kundu, S., Aulchenko, Y. S., van Duijn, C. M. & Janssens, A. C. J. W. PredictABEL: an R package for the assessment of risk prediction models. Eur. J. Epidemiol. 26, 261–264 (2011).

27. Jackson, I. J. MOLECULAR AND DEVELOPMENTAL GENETICS OF MOUSE COAT COLOR. Annual Review of Genetics vol. 28 189–217 Preprint at https://doi.org/10.1146/annurev.ge.28.120194.001201 (1994).

28. Almathen, F., Elbir, H., Bahbahani, H., Mwacharo, J. & Hanotte, O. Polymorphisms in MC1R and ASIP Genes are Associated with Coat Color Variation in the Arabian Camel. J. Hered. 109, 700–706 (2018).

29. Ito, S., Wakamatsu, K. & Sarna, T. Photodegradation of Eumelanin and Pheomelanin and Its Pathophysiological Implications. Photochem. Photobiol. 94, 409–420 (2018).

30. Garrett, G. L. et al. Incidence of and Risk Factors for Skin Cancer in Organ Transplant Recipients in the United States. JAMA Dermatol. 153, 296–303 (2017).

